# Breathing pattern and pulmonary gas exchange in elderly patients with and without left ventricular dysfunction - modification with exercise-based cardiac rehabilitation and prognostic value

**DOI:** 10.1101/2023.03.17.23287430

**Authors:** Prisca Eser, Thimo Marcin, Eva Prescott, Leonie F. Prins, Evelien Kolkman, Wendy Bruins, Astrid E van der Velde, Carlos Peña Gil, Marie-Christine Iliou, Diego Ardissino, Uwe Zeymer, Esther P Meindersma, Arnoud W.J. Van’t Hof, Ed P. de Kluiver, Matthias Wilhelm

## Abstract

**Aims:** In elderly patients with and without left ventricular dysfunction (LVD) we compared ventilatory parameters from before to after exercise-based cardiac rehabilitation (exCR) and assessed their prognostic value for major adverse cardiovascular events (MACE).

**Methods and Results:** Patients aged ≥65 years with acute or chronic coronary syndromes (ACS, CCS) without cardiac surgery who participated in a multicentre study on effectiveness of exCR from seven European countries were included. Cardiopulmonary exercise testing (CPET) was performed before, at termination of exCR and at 12 months follow-up. Ventilation (VE), breathing frequency (BF), tidal volume (VT) and end-expiratory carbon dioxide pressure (P_ET_CO_2_) were measured at rest, at first ventilatory threshold and peak exercise.

In 818 patients, age was 72.5±5.4 years, 21.9% were women, 79.8% had ACS, and 151 (18%) had LVD. NYHA functional class was not different between LVD and noLVD (86% NYHA I in each group). Compared to noLVD, in LVD resting VE was increased 8%, resting BF 6%, peak VE, peak VT, and peak P_ET_CO_2_ reduced by 6%, 8%, and 5%, respectively, and VE/VVCO_2_ slope increased by 11%. From before to after exCR, resting VE decreased and peak P_ET_CO_2_ increased significantly more in LVD compared to noLVD. In LVD, higher resting BF, higher nadir VE/VCO_2_, and lower peak P_ET_CO_2_ at baseline were associated with MACE

**Conclusions:** In elderly patients with ischemic LVD, exaggerated resting ventilation and impaired pulmonary gas exchange during exercise was related to worse outcomes. ExCR was associated with an improvement of abnormal breathing patterns and gas exchange parameters.

## Introduction

Ischemic heart disease is the most prevalent risk factor for left ventricular dysfunction (LVD) and chronic heart failure (HF), both for woman and men.(1) Impaired pulmonary gas exchange, quantified as an increased ventilation (VE) to carbon dioxide exhalation (VCO_2_) slope during exercise, and a low end-tidal pressure of carbon dioxide (P_ET_CO_2_) is a landmark of heart failure patients.(2, 3) These parameters received attention when several studies found higher VE/VCO_2_ slopes and lower P_ET_CO_2_ to be associated with poorer prognosis.(4-6) The components of the VE/VCO_2_ slope are the arterial CO_2_ partial pressure (P_a_CO_2_), that is affected by ventilation, and the pulmonary dead space to tidal volume ratio (VD/VT) that is affected by pulmonary perfusion abnormalities.(3, 7) As nicely summarised by Cross and colleagues, exercise hyperventilation is a hallmark of the failing heart.(2) They proposed several mechanisms as mediators of this excessive ventilatory response, including: 1) alveolar ventilation-perfusion mismatching, 2) increased humoral stimuli (e.g., lactate and H^+^) due to skeletal muscle hypoperfusion and deconditioning, 3) juxta-capillary receptor stimulation consequent to pulmonary vascular congestion and/or hypertension, 4) augmented central and peripheral chemosensitivity, and 5) an inordinately high degree of afferent neural traffic originating from within the locomotor muscles (i.e., the ergoreflex or “skeletal muscle” hypothesis). Hyperventilation is well known to stimulate sympathetic nervous activity.(8) Chronic sympathetic nervous hyperactivity in turn may decrease aerobic capacity of skeletal muscles by reducing capillarisation(9) and red blood cell flux,(10) which lead to a shift in muscle fibre type towards a lower content on type I fibres.(11) The ensuing anaerobic muscle metabolism leads to increased muscle fatiguability(12) and acidosis already at low levels of exercise, which trigger exaggerated responses in ventilation.(13) While exaggerated ventilation has been found to be a strong predictor of mortality and adverse cardiac events in patients with established heart failure, this has not been assessed in patients with LVD.

Comprehensive exercise-based cardiac rehabilitation (exCR) has the potential to improve cardiorespiratory fitness (CRF) and quality of life,(14) cardiovascular risk factors (15), and reduce hospitalisations and cardiovascular mortality.(15, 16) In HFrEF patients, studies assessing the effects of exercise have focussed largely on circulatory parameters,(17) peak oxygen uptake (VO_2_) and muscle strength.(18) Some studies have also reported changes in ventilatory efficiency in response to exercise during exCR.(19, 20) However, data in elderly patients with LVD are sparse and no data exists on how exCR influences breathing patterns at rest.

The purpose of the present study in elderly patients with and without ischemic LVD was to 1) describe breathing patterns and pulmonary gas exchange parameters at rest and during exercise; 2) assess the association of exCR with a change of these parameters; and 3) assess the prognostic value of these parameters on major adverse cardiac events (MACE).

## Methods

The EU-CaRE observational study was a European project focusing on the effectiveness and sustainability of exCR programs in the elderly (65 years or above). EU-CaRE involved eight participating CR sites in seven countries (Denmark, France, Germany, Italy, the Netherlands, Spain and Switzerland).(21)

### Study population

The study population and outcome data have been reported previously.(21-25) For the present study, only data from patients with acute or chronic coronary syndromes (ACS/CCS) with or without percutaneous coronary intervention (PCI) but without open-chest surgery were included. Patients were assessed at baseline before commencing exCR (T0), after completing the exCR program (T1) and at 1-year follow-up (T2). They were grouped according to left ventricular ejection fraction (LVEF) determined by the patient’s most recent echocardiography into patients with LVD (LVEF<45%), versus those without LVD (noLVD, LVEF≥45%). At the time when this study was designed, this cut-off was one of the more widely used and accepted cut-offs for systolic LVD and HF with reduced LVEF (HFrEF).(26)

The study was approved by all relevant medical ethics committees, registered at trialregister.nl (NTR5306). The participants gave written informed consent before they were included in the study.

### Data collection

Recorded information included demographics, index event, socioeconomic factors, medical history including co-morbidity, and clinical information such as weight, blood pressure (BP), resting heart rate (HR), medication, SF36 quality of life questionnaire, and patient reported physical activity as number of days per week with at least moderate physical activity of minimally 30 min. Details on the collected data have been provided elsewhere.(21, 27, 28)

CRF and breathing parameters were assessed by cardiopulmonary exercise testing (CPET). After reporting to the laboratory, patients rested supine for 10-15 min during which a resting 12-lead electrocardiogram (ECG) was performed. Then patients stood up and performed a resting spirometry with determination of forced vital capacity (FVC) and forced expiratory volume during the first second (FEV1). After this, patients mounted the cycle ergometers and were fitted with a facemask. Then they remained sitting quietly for 3 min during which heart rate, blood pressure and breathing parameters were measured. As an average of the last minute of the 3-min resting period while sitting on the ergometer, the following parameters were determined: minute ventilation (VE), breathing frequency (BF), tidal volume (VT), partial pressure of carbon dioxide (P_ET_CO_2_), and heart rate. After a 3-min warm-up at 5 Watt an individual ramp was chosen to achieve a test duration of 8-12 min until exhaustion and kept constant in follow-up tests. Data was analysed at the CPET core lab in Bern as previously described.(24, 28) During the ramp, VE/VCO_2_ slope, and the nadir of the VE/VCO_2_ ratio were determined. Oxygen consumption (VO_2_), VE, BF, VT, P_ET_CO_2_, and heart rate were determined at the first ventilatory thresholdas previously described.(24) Peak values of the same parameters were determined as the highest value of a 30 s moving average. Ventilatory parameters were excluded when respiratory quotient was below 0.7 at resting and below 0.8 at peak exercise due to suspected presence of mask leakage.

MACE was defined as combined incidence of all-cause mortality, ACS, cardiac related emergency visit, hospitalisation for cardiac reason, near sudden cardiac death and cardiac intervention. MACE was recorded by monthly telephone calls and assessed individually by an independent Clinical Event Committee.(24)

### Statistical analysis

All statistics were performed with R (Version 3.5.1, R Core Team, 2017). Changes in ventilatory and circulatory parameters were calculated between T0 and T1. Mixed linear models were performed for ventilatory parameters VE, BF, VT, and P_ET_CO_2_ at rest and peak exercise, as well as VE/VCO_2_ slope and peak VO_2_ relative to body weight for interaction effects between time points T0, T1 and T2 and group (with and without LVD), adjusted for age, sex, and body mass index (BMI) and patients nested within centres entered as random factor (intercepts).

The association of ventilatory parameters at rest and during exercise with MACE were also assessed with mixed linear models for patients with and without LVD and with and without MACE.

## Results

Of 1633 enrolled EU-CaRE patients, 986 patients had an ACS or CCS with percutaneous coronary intervention or no revascularisation. Of these, 79.8% had an ACS and 90.2% a PCI as indication for exCR. LVEF was known in 912 patients. Of these, valid CPET at T0 was available in 867 patients of whom 707 had no LVD and 166 had LVD. Amongst these, data on breathing pattern was available from at least one time point in 151 with and 667 patients without LVD (Figure 1). Baseline data of the two patient groups are shown in Table 1. Patients with LVD were older, were less often female, and had lower BMI and FVC/BSA. An equal prevalence of 86% of patients with NYHA functional class I existed in patients with and without LVD, and NYHA III class was present in 2% and 3% of patients with and without LVD, respectively. Patients with LVD had significantly lower physical component score of the SF36 indicating poorer health status. Patients with LVD had a higher percentage of pre-existing chronic heart failure and anaemia, and higher prescription rate of betablockers and ACE inhibitors or ARBs. With regard to ventilatory parameters, two centres did not record data on BF and VT, therefore, these parameters were missing in 5 and 51 patients with and without LVD, respectively. Due to an insufficient ventilation monitoring duration (≤1 min) during the 3-min resting phase, VE and P_ET_CO_2_ at rest were missing in 9 and 68 patients with and without LV dysfunction at T0, respectively, in 6 and 82 at T1, respectively, and in 4 and 46 at T2, respectively. The first ventilatory threshold could not be determined in 38 and 130 patients with and without LV dysfunction, respectively, at T0, in 20 and 71 at T1, respectively, and in 16 and 61 at T2, respectively. The largest relative adjusted (for sex, age and BMI) differences of the LVD group compared to the noLVD group at baseline were found for VO_2_ peak (−12.6%), VE/VCO_2_ slope (11.0%), nadir VE/VCO_2_ (8.8%), and resting VE (8.1%) (Table 2).

**Table 1:** Baseline characteristics of patients without vs. with LVD. Parameters are indicated as mean ± SD, n (%), or median [1^st^, 3^rd^ quartile].

| Group (n) | No LVD (667) | LVD (151) | p-value |
| --- | --- | --- | --- |
| Age [yrs] | 72.3 ± 5.4 | 73.3 ± 5.4 | 0.0340 |
| Female sex | 156 (23.4) | 21 (13.9) | 0.0145 |
| BMI [kg/m<sup>2</sup>] | 27.7 ± 4.0<sup>c</sup> | 26.8 ± 3.9<sup>a</sup> | 0.0036 |
| Forced vital capacity [l] | 3.31 ± 0.84<sup>c</sup> | 3.17 ± 0.87<sup>a</sup> | 0.0952 |
| Forced vital capacity/BSA [l/m<sup>2</sup>] | 1.14 ± 0.24<sup>c</sup> | 1.08 ± 0.24<sup>a</sup> | 0.0067 |
| Forced expiratory volume in first second [l/s] | 2.59 ± 0.69<sup>c</sup> | 2.49 ± 0.64<sup>a</sup> | 0.0705 |
| *Patient reported outcomes* |  |  |  |
| NYHA class I/II/III | 574/73/17<sup>a</sup> | 130/18/3 | 0.8776 |
| SF36 physical component score | 47.5 [41.1, 52.6]<sup>e</sup> | 44.3 [38.8, 49.4]<sup>d</sup> | 0.0004 |
| Days with >30 min physical activity | 3 [1, 7]<sup>b</sup> | 4 [2, 7]<sup>a</sup> | 0.2366 |
| *Cardiac diseases* |  |  |  |
| Indication for exCR ACS | 516 (77.4) | 129 (85.4) | 0.0373 |
| PCI during index event | 598 (89.7) | 140 (92.7) | 0.3215 |
| Previous ACS | 141 (21.1)<sup>a</sup> | 42 (27.8) | 0.1028 |
| Chronic heart failure | 3 (0.4) | 10 (6.6) | 0.0000 |
| Atrial fibrillation | 33 (4.9) | 12 (7.9) | 0.2069 |
| *CV risk factors* |  |  |  |
| Smoking (active/former/never) | 69/111/485<sup>a</sup> | 14/25/112 | 0.9163 |
| Hypertension | 438 (65.7) | 100 (66.2) | 0.9901 |
| Hypercholesterolemia | 453 (67.9) | 94 (62.3) | 0.1974 |
| Diabetes mellitus | 152 (22.8) | 43 (28.5) | 0.1751 |
| Family history of CVD | 162 (24.3) | 35 (23.2) | 0.9056 |
| *Comorbidities* |  |  |  |
| Nephropathy | 47 (7.0) | 17 (11.3) | 0.1102 |
| Peripheral arterial disease | 44 (6.6) | 12 (7.9) | 0.6630 |
| Chronic obstructive pulmonary disease | 41 (6.1) | 14 (9.3) | 0.2199 |
| Obstructive sleep apnea | 22 (3.3) | 2 (1.3) | 0.3076 |
| Anaemia | 116 (29.0)<sup>f</sup> | 42 (49.4)<sup>g</sup> | 0.0179 |
| *Medication* |  |  |  |
| Beta blockers | 510 (76.5) | 140 (92.7) | 0.0001 |
| Statins | 637 (95.5) | 144 (95.4) | 0.9999 |
| ACE inhibitors or ARBs | 501 (75.1) | 130 (86.1) | 0.0056 |

LVD, left ventricular dysfunction; BMI, body mass index; NYHA, New York Heart Failure Association; SF36, Short-Form Health Survey; CV, cardiovascular; ACS, acute coronary syndrome; CVD, cardiovascular disease; ACE, angiotensin-converting enzyme ; ARB, angiotensin receptor blockers
<sup>a</sup>, missing data $\geq 1$ and $\leq 5$ ; <sup>b</sup>, missing data $\geq 6$ and $\leq 10$ ; <sup>c</sup>, missing data $\geq 11$ and $\leq 15$ ; <sup>d</sup>, missing data = 17; <sup>e</sup>, missing data = 40; <sup>f</sup>, missing data = 151; <sup>g</sup>, missing data = 29

**Table 2:** Ventilatory and circulatory parameters at rest, during ramp exercise (at first ventilatory threshold) and at peak exercise for patients without vs. patients with LVD before start of exCR as well as change until completion of CR. Shown are medians and first and third quartiles in round brackets. Relative difference between groups is derived from group effect from adjusted models and is relative to values from group without LVD. P-values are derived from mixed linear models adjusted for age, sex and body mass index and with patients nested within centres as random intercepts.

| Group (n) | Patients without LVD (667) | Patients with LVD (151) | Relative difference [%] | p-value for group main effect | Change with CR in patients without LVD (609) | Change with CR in patients with LVD (141) | p-value for group x time interaction |
| --- | --- | --- | --- | --- | --- | --- | --- |
| *Resting* |  |  |  |  |  |  |  |
| Ventilation [l/min] | 12.4 (10.2, 14.9) | 13.1 (11.6, 16.2) | **8.1** | **0.0019** | 0.12 (-1.63, 1.83) | -0.93 (-2.96, 1.13) | **0.0018** |
| Breathing frequency [min<sup>-1</sup>] | 17.2 (14.5, 20.5) | 18.0 (14.9, 21.3) | **5.8** | **0.0126** | 0.03 (-2.17, 2.13) | -0.72 (-3.29, 2.03) | 0.1346 |
| Tidal volume [ml/breath] | 736 (570, 895) | 739 (604, 905) | 0.0 | 0.9543 | 1.30 (-133, 141) | -15.1 (-171, 106) | 0.3896 |
| P<sub>ET</sub>CO<sub>2</sub> [mmHg] | 29.2 (26.7, 32.2) | 27.4 (24.6, 30.3) | -2.3 | 0.0537 | 0.14 (-1.68, 1.99) | 0.11 (-1.68, 2.22) | 0.5562 |
| Heart rate [bpm] | 63 (58, 70) | 65 (60, 74) | **3.8** | **0.0245** | -2 (-8, 4) | -3 (-9, 3) | 0.9148 |
| *At first ventilatory threshold* |  |  |  |  |  |  |  |
| VO<sub>2</sub> [ml/kg/min] | 10.9 (9.25, 12.6) | 9.7 (8.6, 11.7) | **-7.5** | **0.0024** | 1.13 (-0.26, 2.83) | 0.92 (-0.50, 2.33) | 0.6841 |
| Ventilation [ml/min] | 341 (288, 403) | 333 (280, 411) | -1.5 | 0.2477 | 37 (-13, 92) | 26 (-29, 80) | 0.6843 |
| Breathing frequency [min<sup>-1</sup>] | 20.5 (18.3, 24.4) | 21.4 (18.3, 24.4) | **5.1** | **0.0441** | 0.57 (-1.37, 2.59) | -0.03 (-2.09, 2.50) | 0.3010 |
| Tidal volume [ml/breath] | 1338 (1126, 1627) | 1239 (998, 1523) | **-6.7** | **0.0120** | 110 (-48, 268) | 65 (-111, 282)} | 0.9715 |
| P<sub>ET</sub>CO<sub>2</sub> [mmHg] | 34.4 (31.7, 37.3) | 32.2 (28.4, 35.1) | **-5.1** | **0.0000** | 0.35 (-1.42, 1.88) | 1.32 (-0.55, 3.25) | 0.0620 |
| *During ramp exercise* |  |  |  |  |  |  |  |
| VE/VCO<sub>2</sub> slope | 32.1 (28.5, 37.2) | 36.6 (31.0, 43.1) | **11.0** | **0.0000** | 0.09 (-3.28, 2.88) | -1.45 (-4.45, 1.56) | 0.3327 |
| Nadir VE/VCO<sub>2</sub> | 33.7 (30.8, 37.7) | 36.7 (33.5, 42.8) | **8.8** | **0.0000** | -0.09 (-1.96, 1.51) | -1.21 (-3.42, 1.00) | **0.0407** |
| *At peak exercise* |  |  |  |  |  |  |  |
| VO<sub>2</sub> [ml/kg/min] | 17.1 (14.1, 20.1) | 15.4 (12.8, 18.0) | **-12.6** | **0.0000** | 1.29 (-0.06, 3.08) | 1.39 (0.05, 2.87) | 0.6422 |
| Ventilation [l/min] | 54.2 (42.7, 68.5) | 53.7 (41.9, 65.0) | **-5.5** | **0.0175** | 5.13 (-0.54, 11.58) | 6.22 (-2.30, 12.15) | 0.6222 |
| Breathing frequency [min<sup>-1</sup>] | 32.5 (28.3, 36.8) | 32.9 (28.7, 37.4) | 1.0 | 0.6166 | 1.75 (-1.18, 4.86) | 1.96 (-0.49, 4.72) | 0.5397 |
| Tidal volume [l/min] | 1.90 (1.54, 2.33) | 1.76 (1.43, 2.18) | **-7.9** | **0.0005** | 0.08 (-0.07, 0.19) | 0.08 (-0.09, 0.21) | 0.3680 |
| Maximal P<sub>ET</sub>CO<sub>2</sub> [mmHg] | 35.7 (32.7, 38.6) | 33.6 (29.7, 36.6) | **-5.4** | **0.0000** | 0.18 (-1.24, 1.76) | 0.97 (-0.50, 2.81) | **0.0055** |
| RER (VCO<sub>2</sub>/VO<sub>2</sub>) | 1.08 (1.01, 1.14) | 1.04 (0.97, 1.12) | **-1.8** | **0.0266** | 0.01 (-0.04, 0.07) | 0.04 (-0.02, 0.10) | **0.0284** |
| Heart rate [bpm] | 114 (103, 130) | 109 (97, 125) | -1.7 | 0.2608 | 2.8 (-3.9, 9.8) | 5.7 (-2.5, 14.0) | 0.1691 |

LVD, left ventricular dysfunction; PetCO2, end-tidal partial pressure of CO2; VO2, oxygen uptake, VE, ventilation; VCO2, carbon dioxide production

**Table 3:** Association of MACE with CPET parameter. Shown are estimates and 95% CI for MACE from the mixed linear models with the respective CPET parameter as dependent variable and covariates age and sex, and with site as random factor. Models were performed for patients with and without LVD and for the total population to test for a group interaction.

|  | LVD (n=151, 25 with MACE) |  | No LVD (n=667, 104 with MACE) |  | Interaction<br>MACE × LVD |
| --- | --- | --- | --- | --- | --- |
|  | Beta | 95%CI | Beta | 95%CI | p-value |
| VO<sub>2</sub> peak [ml/kg/min] | -1.150 | -2.842, 0.520 | -1.249 | -2.136, -0.354 | 0.7608 |
| VE/VCO<sub>2</sub> slope | 3.448 | -0.092, 7.046 | 2.244 | 0.757, 3.739 | 0.3159 |
| Nadir VE/VCO<sub>2</sub> | 2.843 | 0.071, 5.615 | 1.189 | 0.091, 2.294 | 0.1022 |
| P<sub>ET</sub>CO<sub>2</sub> resting [mmHg] | -0.951 | -2.573, 0.653 | -0.250 | -1.093, 0.593 | 0.3158 |
| P<sub>ET</sub>CO<sub>2</sub> peak [mmHg] | -1.880 | -3.759, -0.019 | -0.404 | -1.328, 0.515 | 0.0860 |
| VE resting [l/min] | 0.882 | -0.853, 2.632 | -0.795 | -1.613, 0.027 | 0.0742 |
| VE peak [l/min] | 2.644 | -3.855, 9.105 | -2.302 | -5.390, 0.803 | 0.3411 |
| BF resting [min<sup>-1</sup>] | 2.985 | 0.639, 5.418 | -0.688 | -2.097, 0.714 | 0.0086 |
| BF peak [min<sup>-1</sup>] | 2.928 | -0.265, 6.122 | 1.200 | -0.506, 2.888 | 0.5638 |
| VT resting [l] | -0.055 | -0.165, 0.054 | -0.018 | -0.073, 0.038 | 0.4746 |
| VT peak [l] | -0.155 | -0.334, 0.021 | -0.075 | -0.173, 0.023 | 0.2658 |

LVD, left ventricular dysfunction; MACE, major adverse cardiac event; VO<sub>2</sub>, oxygen uptake; VE/VCO<sub>2</sub>, ventilation/carbon dioxide production; P<sub>ET</sub>CO<sub>2</sub>, endtidal carbon dioxide partial pressure; BF, breathing frequency; VT, tidal volume

**Figure 1:**
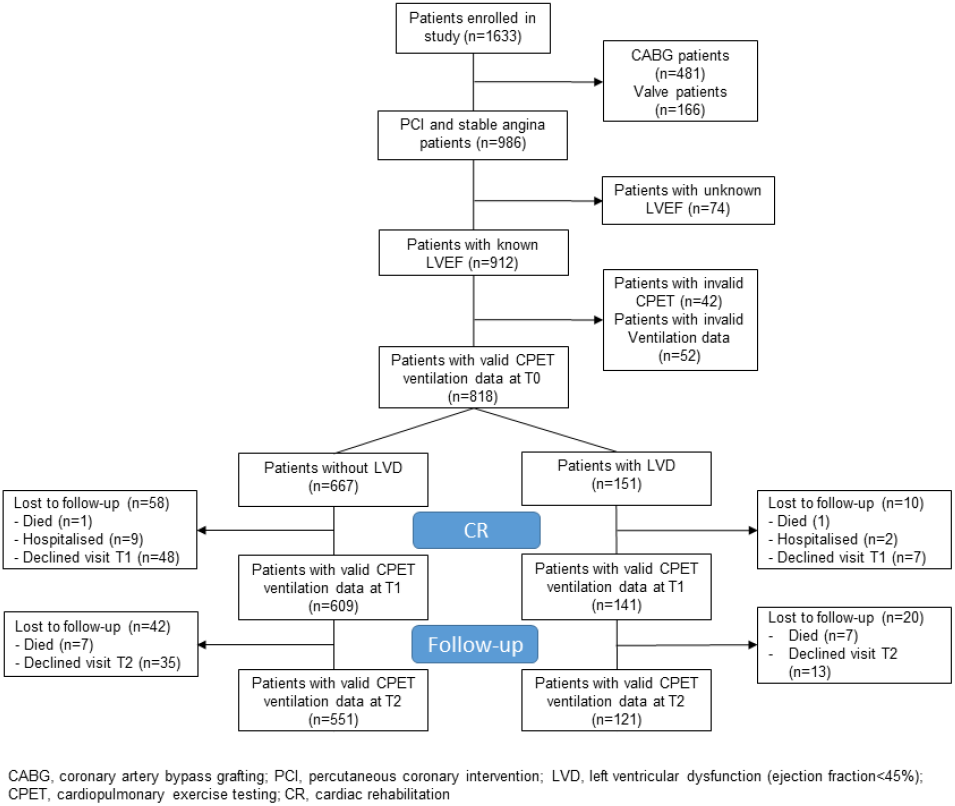
Study flow.

Regarding the changes in ventilatory parameters from before to after CR, patients with LVD had significantly greater reductions in resting VE (Table 2). They also had greater reductions in nadir VE/VCO_2_ (graphical abstract), and a greater increase in P_ET_CO_2_ and RER at peak exercise.

Results from the mixed linear models including data also for the 1-year follow-up measurement as well as confounding factors age, sex and BMI showed a significantly greater reduction of VE at rest in the group of patients with LVD not only from before to after exCR but also to 1-year follow-up (Figure 2, Supplement Table 1a). BF at rest was higher in patients with LVD at all time points (Figure 2, Supplement Table 1a), and resting P_ET_CO_2_ increased significantly at 1-year follow-up in both groups (Figure 3, Supplement Table 1a). At peak exercise, VE was reduced in the patients with LVD due to a significantly lower VT in the patients with LVD but increased with exCR and to 1-year follow-up in both groups (Figure 2, Supplement Table 1b),. Resting P_ET_CO_2_ was lower by trend (p=0.0537) in patients with LVD and significantly lower at peak exercise at all time points (Figure 3, Supplement Table 1a and b). Accordingly, VE/VCO_2_ slope was increased in patients with LVD at all time points (Figure 3, Supplement Table 1c). VO_2_ relative to body weight was greatly reduced in patients with LVD at peak exercise (Figure 3, Supplement Table 1c) at all time points. Peak VO_2_ increased in both groups from before to after exCR and VE/VCO_2_ slope decreased over time. The physical component score was consistently lower by 3 points in patients with LVD but significant consistent improvements in SF36 PCS between time points were comparable in both groups with nearly 3 points between T0 and T2 (Supplement Table 1c).

**Figure 2:**
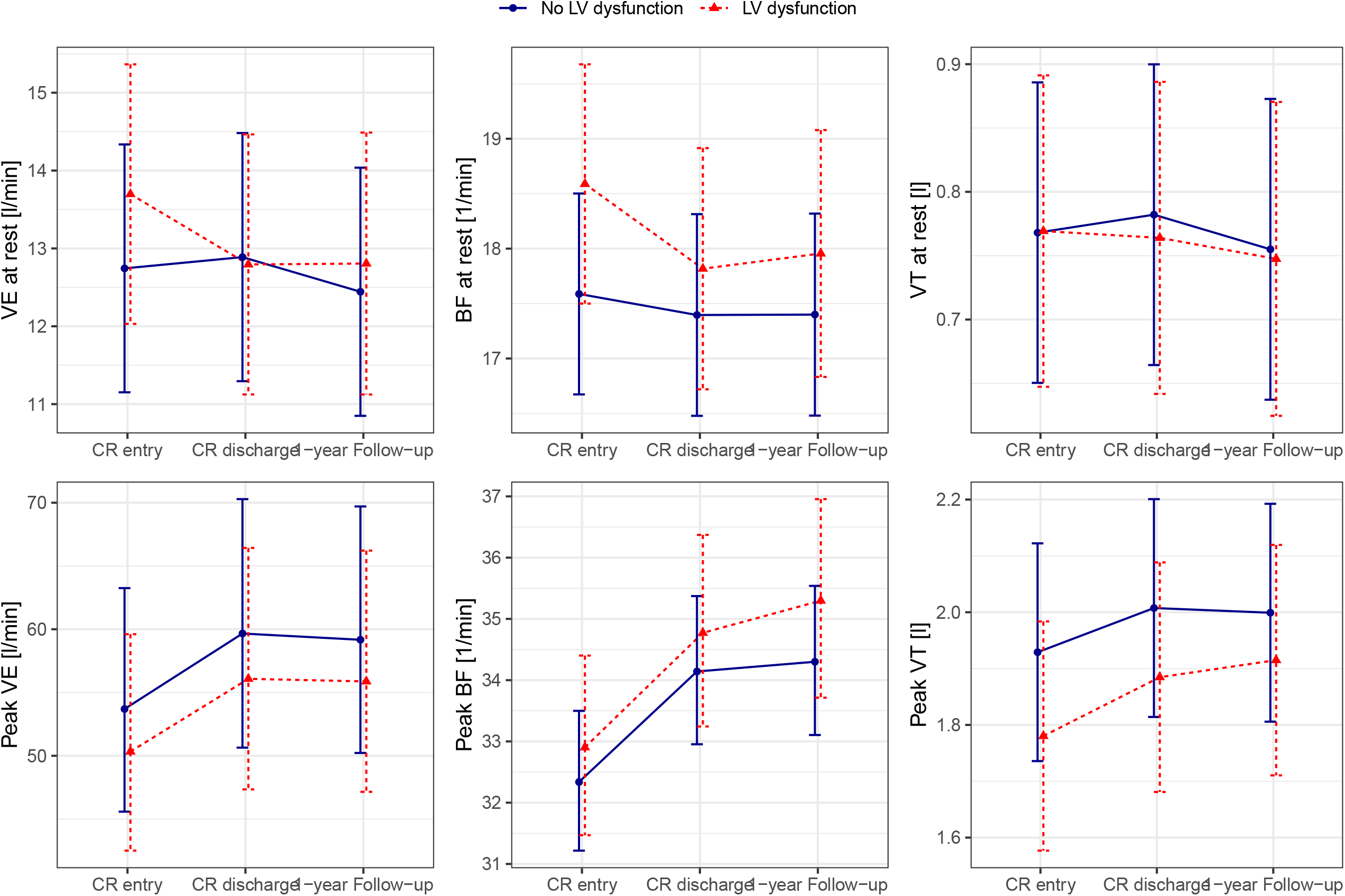
Predicted means with 95% confidence intervals based on the mixed linear models for ventilation (a), breathing frequency (b), and tidal volume (c) at rest and at peak exercise (d-f) adjusted for age, sex, and body mass index, with patients nested within centres as random intercepts.

**Figure 3:**
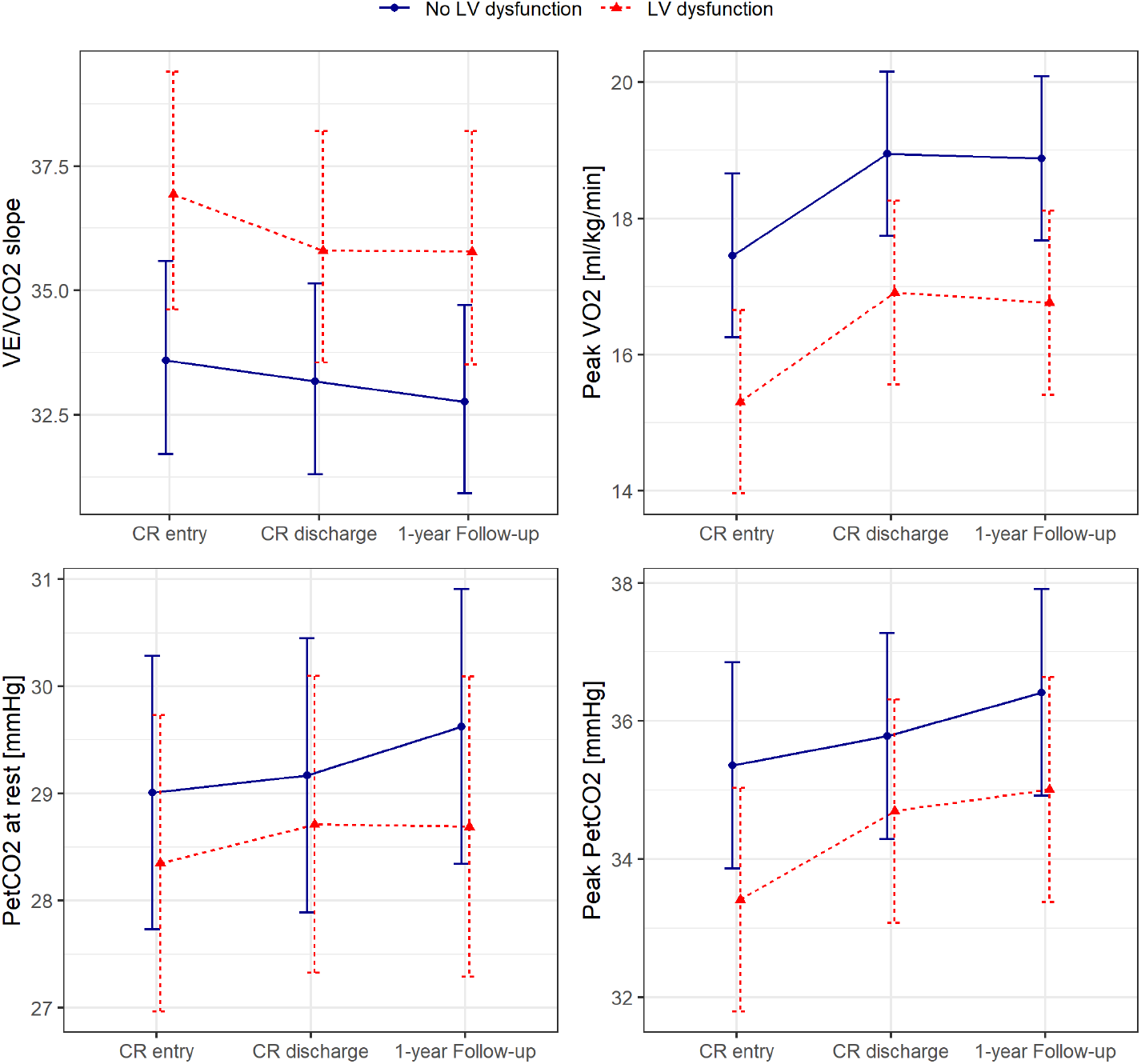
Predicted means with 95% confidence intervals based on the mixed linear models for VE/VCO_2_ slope (a), peak VO_2_ (b), and end-tidal carbon dioxide partial pressure at rest (c) and at peak exercise (d) adjusted for age, sex, and body mass index, with patients nested within centres as random intercepts.

Approximately 20% of patients with and without LVD had a MACE within 1-year follow-up with no difference between groups (p-value for Chi-square test 0.069). Nadir of VE/VCO_2_ significantly discriminated patients with and without MACE in patients with and without LVD (graphical abstract), while VE/VCO_2_ slope and peak VO_2_ discriminated for MACE only in patients without LVD, and peak P_ET_CO_2_ and resting BF discriminated for MACE only in patients with LVD.

## Discussion

This study provides four clinically relevant and novel aspects in a representative European cohort of elderly patients with coronary artery disease undergoing exCR: 1) Nearly one fifth of patients had asymptomatic LVD at the start of exCR; 2) LVD was associated with elevated ventilation at rest and an impaired pulmonary gas exchange during exercise; 3) exCR was associated with an improvement of ventilation at rest and gas exchange parameters during exercise, but ventilatory efficiency and P_ET_CO2 remained significantly reduced in patients with LVD, compared to patient without LVD in the 1-year follow-up; and 4) BF at rest, ventilatory efficiency and P_ET_CO_2_ during exercise were associated with worse outcome in patients with LVD.

This is the first large study demonstrating an increased resting ventilation and impaired pulmonary gas exchange during exercise in patients with LVD without major signs and symptoms of congestive heart failure. NYHA functional class did not differ between patients with and without LVD and 86% of patients in both groups were in NYHA functional class I. Furthermore, self-reported physical activity did not differ between the groups. Although the median physical component score of the SF36 questionnaire was 3 points lower in patients with LVD compared to patients without LVD, the values of 44.3 at T0 and 47.5 at T2 were clearly above the median of 35.3 found in patients with chronic heart failure.(29)

We suggest that signs of an exaggerated ventilatory drive are already apparent in patients with LVD at rest (higher resting VE and BF), during exercise (increased VE/VCO_2_ slope and nadir VE/VCO_2_), and at peak exercise (lower VO_2_, VT and P_ET_CO_2_). Reductions of the exaggerated ventilatory drive seen with exCR were similarly beneficial in patients with and without LVD. The decrease in nadir VE/VCO_2_ was significantly greater in patients with LVD than in those without LVD, with similar magnitude of the difference between changes in VE/VCO_2_ slope, albeit not reaching statistical significance. This is in accordance to some previous studies that found ventilatory efficiency to improve most in patients with most severe ventilatory inefficiency at baseline. For example, in a study with 131 patients after acute myocardial infarction without HF participating in exCR, VE/VCO_2_ slope improved most in patients with VE/VCO_2_ slope>32,(30) which was also confirmed by a study comparing the effect of aerobic training to a combination of aerobic and strength training.(31) Similarly, a study in patients with HFrEF by Servantes et al. found large decreases in VE/VCO_2_ slopes with mean pre-exercise values of 36.(32) In part, a larger decrease in patients with higher starting values may be due to a phenomenon referred to as “regression to the mean”.(33) In a study including 123 CAD patients, patients in the group with peak VO_2_<17.5 ml/kg/min also had the highest nadir VE/VCO_2_.(34) This group also had the highest improvement in peak VO_2_ and nadir VE/VCO_2_ after exCR. Likewise, in 37 CAD patients completing exCR, P_ET_CO_2_ at the first and second ventilatory threshold and peak exercise were improved after the programme. On the contrary, Fu et al. did not demonstrate an improvement of the VE/VCO_2_ slope with exercise training in patients with HFrEF and VE/VCO_2_ slope of 35 at baseline.(19)

At peak exercise, peak ventilation was 3 l/min (6%) less in patients with compared to patients without LVD, while peak VO_2_ was reduced by 2.2 ml/kg/min (12%). The disproportionate reduction in gas exchange was likely to be due to the significantly smaller peak VT (150 ml or 8%) in patients with LVD at an insignificantly increased peak BF, resulting in a larger anatomical dead space. This is in line with observation in patients with HFrEF,(35) and the emerging evidence that HF patients adopt a “rapid shallow” breathing pattern to avoid the adverse effects of large intrathoracic pressure swings on cardiac pre- and afterload.(2) Importantly, our study extends the evidence on abnormal breathing patterns and hyperventilation in patients with HFrEF to patients with LVD. The significantly higher resting HR in patients with LVD compared to patients without LVD is compatible with sympathetic hyperactivity.(2, 36) This finding suggests that several proposed mechanisms for exaggerated ventilation in HFrEF (e.g. augmented peripheral and central chemosensitivity and an inordinately high degree of afferent neural traffic originating from within the locomotors muscles) may already be present in patients with asymptomatic LVD starting the vicious cycle that may progress LVD to HFrEF.(2) Interestingly, the models corrected for age, sex and BMI showed a significant positive relationship of BMI with P_ET_CO_2_ at rest and at AT, and a significant inverse relationship with nadir VE/VCO_2_. This raises the question whether the obesity paradox not only applies to patients with HF but already to elderly patients with (and without) LVD.(37)

The observed normalisation of the excessive ventilation of patients with LVD after exCR may partly be ascribed to the beneficial effects of exercise training on ergoreflex sensitivity, and/or chemoreceptor activation.(36) Importantly, guideline directed medical and device therapies for HFrEF improve chemo- and baroreceptor function insufficiently, highlighting the importance of exercise in this population.(38)

While the VE/VCO_2_ slope is an established prognostic parameter in patients with HFrEF, only few studies have reported the physiologically inversely related parameter P_ET_CO_2_ at rest and at peak exercise.(39, 40) Matsumoto and colleagues have found severity of HF in 112 patients with cardiac disease to be negatively associated with P_ET_CO_2_ at rest and at peak exercise.(39) They found P_ET_CO_2_ at peak exercise to be negatively associated with VE/VCO_2_ slope and VD/VT, but not with P_a_CO_2_. Arena and colleagues found a resting P_ET_CO_2_ of <33 mmHg to discriminate significantly for cardiac events at 1-year follow-up and to significantly add discriminative power to VE/VCO_2_ slope.(40) Our study demonstrated consistent findings on the prognostic value of pulmonary gas exchange parameters in patients with LVD. The nadir VE/VCO_2_ and peak P_ET_CO_2_ were associated with MACE. Moreover, as a novel finding, BF at rest was associated with MACE. The discriminative value of resting ventilation parameters for MACE deserves further investigation as both resting P_ET_CO_2_ and BF have been found to be predictive for cardiovascular complications also in lung resection patients.(41)

The important role of low P_ET_CO_2_ in patients with HFrEF and periodic breathing has been pointed out by a study by Apostolo and colleagues who restored a normal breathing pattern with inhalation of 2% CO_2_ in 95% of their study patients.(42) Likewise, inhalation of CO_2_ has been shown to stabilise breathing in patients with periodic breathing and Cheyne-Stokes respiration during sleep.(43-45) Breathing instabilities are recognised markers for poor prognosis in patients with HFrEF and highlight the central role of breathing patterns in the progression of HF.(46) Nevertheless, few therapies have been developed to address breathing patterns. A few small studies with encouraging results have employed slow breathing training in patients with heart failure.(47, 48)

Strengths of this study are the large sample size of a representative European cohort of elderly patients with coronary artery disease undergoing exCR, and the longitudinal as well as comprehensive assessment of ventilatory parameters at rest, during ramp exercise and at peak exercise. Furthermore, this is the first documentation of improved breathing pattern and pulmonary gas exchange both at rest and with exercise after completion of exCR in both patient with and without LVD.

This explorative study is limited by multiple testing. Therefore, confidence intervals and p-values have to be interpreted accordingly. Unfortunately, we did not have data on betablocker classes. Previous studies showed that carvedilol, an unselective ß1- and ß2-blocker, was associated with a lower VE/VCO_2_ slope and nadir VE/VCO_2_.(49, 50) However, standard treatment of ACS and CCS includes cardioselective betablockers like metoprolol. We can therefore only speculate that only a small proportion of the 93% of our patients have received an unselective betablocker. Further, ACE inhibitors have been shown to improve gas transfer and ventilation-perfusion.(51) Nevertheless, the higher percentage of ACE in patients with LVD could not completely offset their less efficient ventilation.

Logistic regressions for identification of CPET variables significantly improving MACE prediction were performed for the whole EU-CaRE cohort (including patients with open heart surgery and valve replacement) recently.(24) Due to the small sample size of our subgroup of patients with LVD, we did not perform logistic regression models in the present study.

## Conclusion

Patients with LVD had an exaggerated BF at rest and an impaired pulmonary gas exchange during exercise. Abnormal breathing patterns may be an early and clinically relevant sign of LVD and linked to increased ergoreflex sensitivity and/or abnormal chemosensitivity. ExCR may contribute to improvements of breathing patterns and pulmonary gas exchange in this population. However, interventions aimed at specifically improving altered breathing patterns at rest and during exercise may have additive value and should be investigated.

## Data Availability

Data may be available upon request.

## Acknowledgements

We would like to thank Judith Peterhans for her valuable contributions to study logistics and data collection, and Susana Pérez-Alves for the graphical abstract design.

---

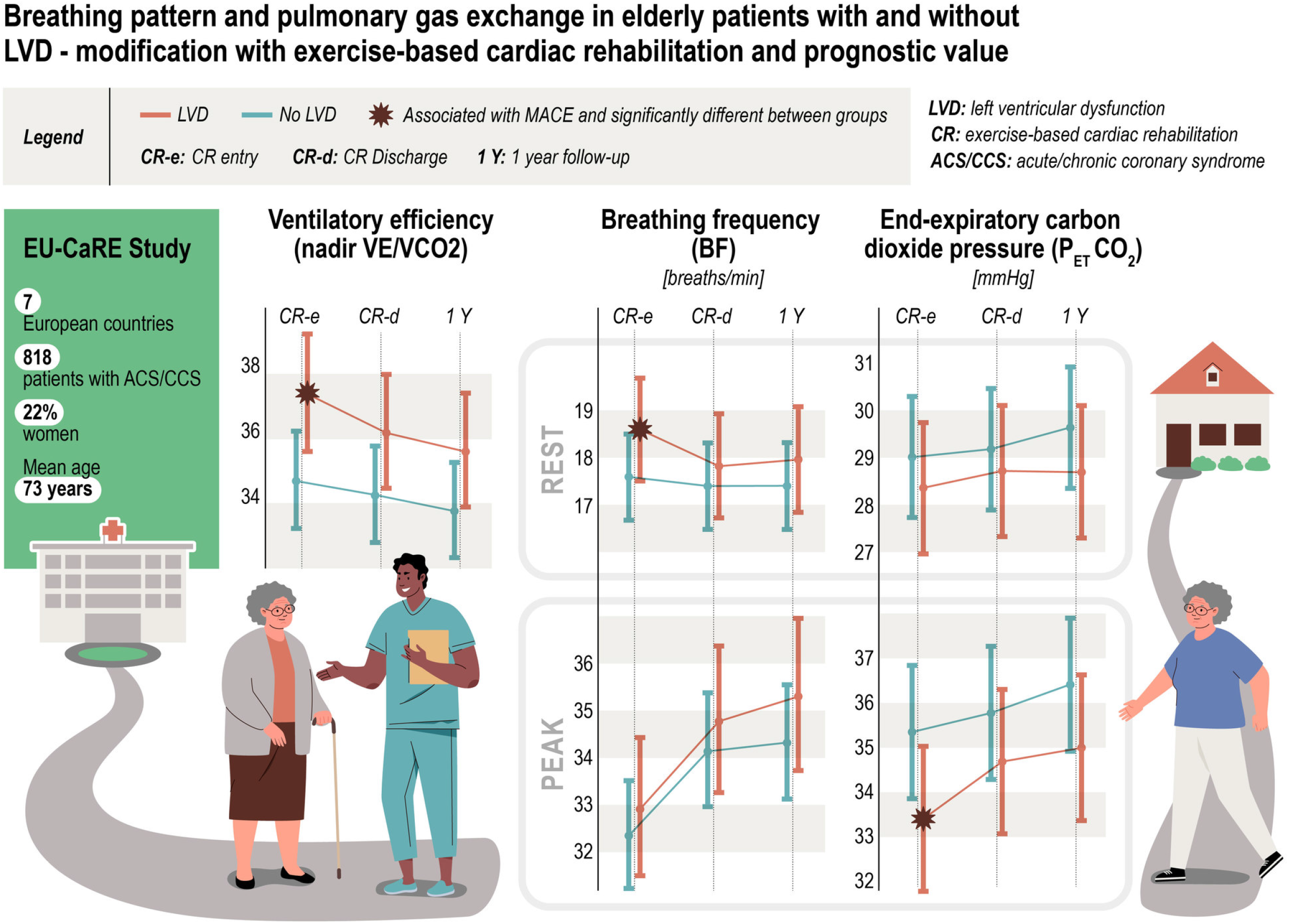

## References

1. Lawson CA, Zaccardi F, Squire I, Okhai H, Davies M, Huang W, et al. Risk Factors for Heart Failure: 20-Year Population-Based Trends by Sex, Socioeconomic Status, and Ethnicity. Circulation Heart failure. 2020;13(2):e006472.

2. Cross TJ, Kim CH, Johnson BD, Lalande S. The interactions between respiratory and cardiovascular systems in systolic heart failure. J Appl Physiol (1985). 2020;128(1):214–24.

3. Malhotra R, Bakken K, D’Elia E, Lewis GD. Cardiopulmonary Exercise Testing in Heart Failure. JACC Heart Fail. 2016;4(8):607–16.

4. Ponikowski P, Francis DP, Piepoli MF, Davies LC, Chua TP, Davos CH, et al. Enhanced ventilatory response to exercise in patients with chronic heart failure and preserved exercise tolerance: marker of abnormal cardiorespiratory reflex control and predictor of poor prognosis. Circulation. 2001;103(7):967–72.

5. Myers J, Arena R, Oliveira RB, Bensimhon D, Hsu L, Chase P, et al. The lowest VE/VCO2 ratio during exercise as a predictor of outcomes in patients with heart failure. Journal of cardiac failure. 2009;15(9):756–62.

6. Nadruz W, Jr., West E, Sengelov M, Santos M, Groarke JD, Forman DE, et al. Prognostic Value of Cardiopulmonary Exercise Testing in Heart Failure With Reduced, Midrange, and Preserved Ejection Fraction. Journal of the American Heart Association. 2017;6(11).

7. Johnson RL, Jr. Gas exchange efficiency in congestive heart failure. Circulation. 2000;101(24):2774–6.

8. Coats AJ, Clark AL, Piepoli M, Volterrani M, Poole-Wilson PA. Symptoms and quality of life in heart failure: the muscle hypothesis. Br Heart J. 1994;72(2 Suppl):S36–9.

9. Duscha BD, Kraus WE, Keteyian SJ, Sullivan MJ, Green HJ, Schachat FH, et al. Capillary density of skeletal muscle: a contributing mechanism for exercise intolerance in class II-III chronic heart failure independent of other peripheral alterations. Journal of the American College of Cardiology. 1999;33(7):1956–63.

10. Hirai DM, Musch TI, Poole DC. Exercise training in chronic heart failure: improving skeletal muscle O2 transport and utilization. Am J Physiol Heart Circ Physiol. 2015;309(9):H1419–39.

11. Sullivan MJ, Duscha BD, Klitgaard H, Kraus WE, Cobb FR, Saltin B. Altered expression of myosin heavy chain in human skeletal muscle in chronic heart failure. Med Sci Sports Exerc. 1997;29(7):860–6.

12. Schulze PC, Linke A, Schoene N, Winkler SM, Adams V, Conradi S, et al. Functional and morphological skeletal muscle abnormalities correlate with reduced electromyographic activity in chronic heart failure. Eur J Cardiovasc Prev Rehabil. 2004;11(2):155–61.

13. Piepoli M, Clark AL, Volterrani M, Adamopoulos S, Sleight P, Coats AJ. Contribution of muscle afferents to the hemodynamic, autonomic, and ventilatory responses to exercise in patients with chronic heart failure: effects of physical training. Circulation. 1996;93(5):940–52.

14. Mitchell BL, Lock MJ, Davison K, Parfitt G, Buckley JP, Eston RG. What is the effect of aerobic exercise intensity on cardiorespiratory fitness in those undergoing cardiac rehabilitation? A systematic review with meta-analysis. Br J Sports Med. 2019;53(21):1341–51.

15. Doimo S, Fabris E, Piepoli M, Barbati G, Antonini-Canterin F, Bernardi G, et al. Impact of ambulatory cardiac rehabilitation on cardiovascular outcomes: a long-term follow-up study. European heart journal. 2019;40(8):678–85.

16. Anderson L, Oldridge N, Thompson DR, Zwisler AD, Rees K, Martin N, et al. Exercise-Based Cardiac Rehabilitation for Coronary Heart Disease: Cochrane Systematic Review and Meta-Analysis. Journal of the American College of Cardiology. 2016;67(1):1–12.

17. Haykowsky MJ, Liang Y, Pechter D, Jones LW, McAlister FA, Clark AM. A meta-analysis of the effect of exercise training on left ventricular remodeling in heart failure patients: the benefit depends on the type of training performed. Journal of the American College of Cardiology. 2007;49(24):2329–36.

18. Ruku DM, Tran Thi TH, Chen HM. Effect of center-based or home-based resistance training on muscle strength and VO(2) peak in patients with HFrEF: A systematic review and meta-analysis. Enferm Clin (Engl Ed). 2021.

19. Fu TC, Yang NI, Wang CH, Cherng WJ, Chou SL, Pan TL, et al. Aerobic Interval Training Elicits Different Hemodynamic Adaptations Between Heart Failure Patients with Preserved and Reduced Ejection Fraction. Am J Phys Med Rehabil. 2016;95(1):15–27.

20. Coats AJ, Adamopoulos S, Radaelli A, McCance A, Meyer TE, Bernardi L, et al. Controlled trial of physical training in chronic heart failure. Exercise performance, hemodynamics, ventilation, and autonomic function. Circulation. 1992;85(6):2119–31.

21. Prescott E, Eser P, Mikkelsen N, Holdgaard A, Marcin T, Wilhelm M, et al. Cardiac rehabilitation of elderly patients in eight rehabilitation units in western Europe: Outcome data from the EU-CaRE multicentre observational study. Eur J Prev Cardiol. 2020;27(16):1716–29.

22. Eser P, Marcin T, Prescott E, Prins LF, Kolkman E, Bruins W, et al. Clinical outcomes after cardiac rehabilitation in elderly patients with and without diabetes mellitus: The EU-CaRE multicenter cohort study. Cardiovasc Diabetol. 2020;19(1):37.

23. Marcin T, Eser P, Prescott E, Prins LF, Kolkman E, Bruins W, et al. Training intensity and improvements in exercise capacity in elderly patients undergoing European cardiac rehabilitation - the EU-CaRE multicenter cohort study. PloS one. 2020;15(11):e0242503.

24. Marcin T, Eser P, Prescott E, Prins LF, Kolkman E, Bruins W, et al. Changes and prognostic value of cardiopulmonary exercise testing parameters in elderly patients undergoing cardiac rehabilitation: The EU-CaRE observational study. PloS one. 2021;16(8):e0255477.

25. Eser P, Marcin T, Prescott E, Prins LF, Kolkman E, Bruins W, et al. Predictors for one-year outcomes of cardiorespiratory fitness and cardiovascular risk factor control after cardiac rehabilitation in elderly patients: The EU-CaRE study. PloS one. 2021;16(8):e0255472.

26. Kelly JP, Mentz RJ, Mebazaa A, Voors AA, Butler J, Roessig L, et al. Patient selection in heart failure with preserved ejection fraction clinical trials. Journal of the American College of Cardiology. 2015;65(16):1668–82.

27. Prescott E, Mikkelsen N, Holdgaard A, Eser P, Marcin T, Wilhelm M, et al. Cardiac rehabilitation in the elderly patient in eight rehabilitation units in Western Europe: Baseline data from the EU-CaRE multicentre observational study. Eur J Prev Cardiol. 2019;26(10):1052–63.

28. Marcin T, Eser P, Prescott E, Mikkelsen N, Prins LF, Kolkman EK, et al. Predictors of pre-rehabilitation exercise capacity in elderly European cardiac patients - The EU-CaRE study. Eur J Prev Cardiol. 2020;27(16):1702–12.

29. Huber A, Oldridge N, Höfer S. International SF-36 reference values in patients with ischemic heart disease. Qual Life Res. 2016;25(11):2787–98.

30. Satoh T, Okano Y, Takaki H, Matsumoto T, Yasumura Y, Aihara N, et al. Excessive ventilation after acute myocardial infarction and its improvement in 4 months. Jpn Circ J. 2001;65(5):399–403.

31. Laoutaris ID, Adamopoulos S, Manginas A, Panagiotakos DB, Kallistratos MS, Doulaptsis C, et al. Benefits of combined aerobic/resistance/inspiratory training in patients with chronic heart failure. A complete exercise model? A prospective randomised study. International journal of cardiology. 2013;167(5):1967–72.

32. Servantes DM, Pelcerman A, Salvetti XM, Salles AF, de Albuquerque PF, de Salles FC, et al. Effects of home-based exercise training for patients with chronic heart failure and sleep apnoea: a randomized comparison of two different programmes. Clin Rehabil. 2012;26(1):45–57.

33. Stigler SM. Regression towards the mean, historically considered. Stat Methods Med Res. 1997;6(2):103–14.

34. Prado DM, Rocco EA, Silva AG, Silva PF, Lazzari JM, Assumpção GL, et al. The influence of aerobic fitness status on ventilatory efficiency in patients with coronary artery disease. Clinics (Sao Paulo). 2015;70(1):46–51.

35. Witte KK, Thackray SD, Nikitin NP, Cleland JG, Clark AL. Pattern of ventilation during exercise in chronic heart failure. Heart. 2003;89(6):610–4.

36. Aimo A, Saccaro LF, Borrelli C, Fabiani I, Gentile F, Passino C, et al. The ergoreflex: how the skeletal muscle modulates ventilation and cardiovascular function in health and disease. European journal of heart failure. 2021;23(9):1458–67.

37. Lavie CJ, De Schutter A, Alpert MA, Mehra MR, Milani RV, Ventura HO. Obesity paradox, cachexia, frailty, and heart failure. Heart Fail Clin. 2014;10(2):319–26.

38. Giannoni A, Gentile F, Buoncristiani F, Borrelli C, Sciarrone P, Spiesshoefer J, et al. Chemoreflex and Baroreflex Sensitivity Hold a Strong Prognostic Value in Chronic Heart Failure. JACC Heart Fail. 2022.

39. Matsumoto A, Itoh H, Eto Y, Kobayashi T, Kato M, Omata M, et al. End-tidal CO2 pressure decreases during exercise in cardiac patients: association with severity of heart failure and cardiac output reserve. Journal of the American College of Cardiology. 2000;36(1):242–9.

40. Arena R, Peberdy MA, Myers J, Guazzi M, Tevald M. Prognostic value of resting end-tidal carbon dioxide in patients with heart failure. International journal of cardiology. 2006;109(3):351–8.

41. Mazur A, Brat K, Homolka P, Merta Z, Svoboda M, Bratova M, et al. Ventilatory efficiency is superior to peak oxygen uptake for prediction of lung resection cardiovascular complications. PloS one. 2022;17(8):e0272984.

42. Apostolo A, Agostoni P, Contini M, Antonioli L, Swenson ER. Acetazolamide and inhaled carbon dioxide reduce periodic breathing during exercise in patients with chronic heart failure. Journal of cardiac failure. 2014;20(4):278–88.

43. Lorenzi-Filho G, Rankin F, Bies I, Douglas Bradley T. Effects of inhaled carbon dioxide and oxygen on cheyne-stokes respiration in patients with heart failure. Am J Respir Crit Care Med. 1999;159(5 Pt 1):1490–8.

44. Steens RD, Millar TW, Su X, Biberdorf D, Buckle P, Ahmed M, et al. Effect of inhaled 3% CO2 on Cheyne-Stokes respiration in congestive heart failure. Sleep. 1994;17(1):61–8.

45. Giannoni A, Baruah R, Willson K, Mebrate Y, Mayet J, Emdin M, et al. Real-time dynamic carbon dioxide administration: a novel treatment strategy for stabilization of periodic breathing with potential application to central sleep apnea. Journal of the American College of Cardiology. 2010;56(22):1832–7.

46. Giannoni A, Emdin M, Bramanti F, Iudice G, Francis DP, Barsotti A, et al. Combined increased chemosensitivity to hypoxia and hypercapnia as a prognosticator in heart failure. Journal of the American College of Cardiology. 2009;53(21):1975–80.

47. Parati G, Malfatto G, Boarin S, Branzi G, Caldara G, Giglio A, et al. Device-guided paced breathing in the home setting: effects on exercise capacity, pulmonary and ventricular function in patients with chronic heart failure: a pilot study. Circulation Heart failure. 2008;1(3):178–83.

48. Lachowska K, Bellwon J, Narkiewicz K, Gruchała M, Hering D. Long-term effects of device-guided slow breathing in stable heart failure patients with reduced ejection fraction. Clinical research in cardiology : official journal of the German Cardiac Society. 2019;108(1):48–60.

49. Agostoni P, Apostolo A, Cattadori G, Salvioni E, Berna G, Antonioli L, et al. Effects of beta-blockers on ventilation efficiency in heart failure. American heart journal. 2010;159(6):1067–73.

50. Contini M, Apostolo A, Cattadori G, Paolillo S, Iorio A, Bertella E, et al. Multiparametric comparison of CARvedilol, vs. NEbivolol, vs. BIsoprolol in moderate heart failure: the CARNEBI trial. International journal of cardiology. 2013;168(3):2134–40.

51. Guazzi M, Melzi G, Marenzi GC, Agostoni P. Angiotensin-converting enzyme inhibition facilitates alveolar-capillary gas transfer and improves ventilation-perfusion coupling in patients with left ventricular dysfunction. Clin Pharmacol Ther. 1999;65(3):319–27.

